# Socio-demographic and Clinical Characteristics of Adults with SARS-CoV-2 Infection in Two Hospitals in Bogota, Colombia

**DOI:** 10.1101/2020.08.13.20167445

**Authors:** Alejandro Moscoso D, Alejandra Sanchez, Adriana Aya, Carolina Gomez, Yazmin Rodriguez, Javier Garzon, Felipe Lobelo

**Author notes:** Address correspondence to: Alejandro Moscoso, MD, Msc Research Coordinator, Clinica del Country & Clinica la Colina.

## Abstract

This study is a retrospective cohort of 122 adult patients with SARS-CoV-2 diagnosed and managed at two medium-sized, tertiary private Hospitals. The analysis includes demographic and socio-economic information, symptoms, comorbidities, laboratory test results, therapeutic management, clinical outcomes and complications.

## INTRODUCTION

As of May 29^th^, 2020, there were over 2.6 million Coronavirus disease 2019 (COVID-19) cases and over 151 thousand deaths in the Americas. Surpassing the U.S. and Europe, most new cases and deaths are now being reported in Latin America (LA), the new epicenter of the pandemic.^1^ The first confirmed case of severe acute respiratory syndrome coronavirus 2 (SARS-CoV-2) was reported in Colombia on March 6, 2020. By March 25^th^ the government instituted a national lockdown. We report the characteristics of patients diagnosed and managed at Clinica del Country (CC) and Clinica la Colina (CLC), two medium-sized, tertiary private Hospitals, located in the northern region of Bogota, the fourth largest urban center in LA with a population of 7.86 million.

## METHODS

This study is a retrospective cohort of 122 adult patients (≥18 years) with positive polymerase chain reaction tests for SARS-CoV-2, seen at CC & CLC from March 12 to May 25, 2020. Testing, triaging and clinical management procedures were adopted from national guidelines.^2^

Analyses include demographic and socio-economic (SES) information, symptoms, comorbidities, laboratory test results, therapeutic management, clinical outcomes and complications. Data was stratified by ambulatory care (AC), general ward hospitalization (GW) and Intensive care unit (ICU). Two of 27 ICU-managed patients were referred to other hospitals and their outcomes unknown.

We report numbers (percentages) for categorical variables and medians (interquartile ranges) for continuous variables. In-hospital mortality and discharge dispositions are reported as of May 29, with 5.7% of the cohort still hospitalized. The CC institutional review board approved the project with a waiver of informed consent.

## RESULTS

Of 2010 tests performed, 122 (6%) were positive for SARS-CoV-2. Of these, 26 (21.3%) were managed in the GW and 27 (22%) in the ICU. The cohort median age was 46 years (33.8-57.3), 70 (57.4%) were women and 43 (35.2%) health workers; (Table1). Most patients had private health insurance 93(76.2%) and lived in northeast neighborhoods. (https://bit.ly/36VE9Sm)

The majority of patients presented with cough, fever and shortness of breath and 29 (23,8%) had diarrhea/vomiting. Overall, the most prevalent comorbidities were hypertension and metabolic diseases. Most patients requiring hospitalization were in the 45-64 years group (22%). (Figure 1)

**Figure 1.**
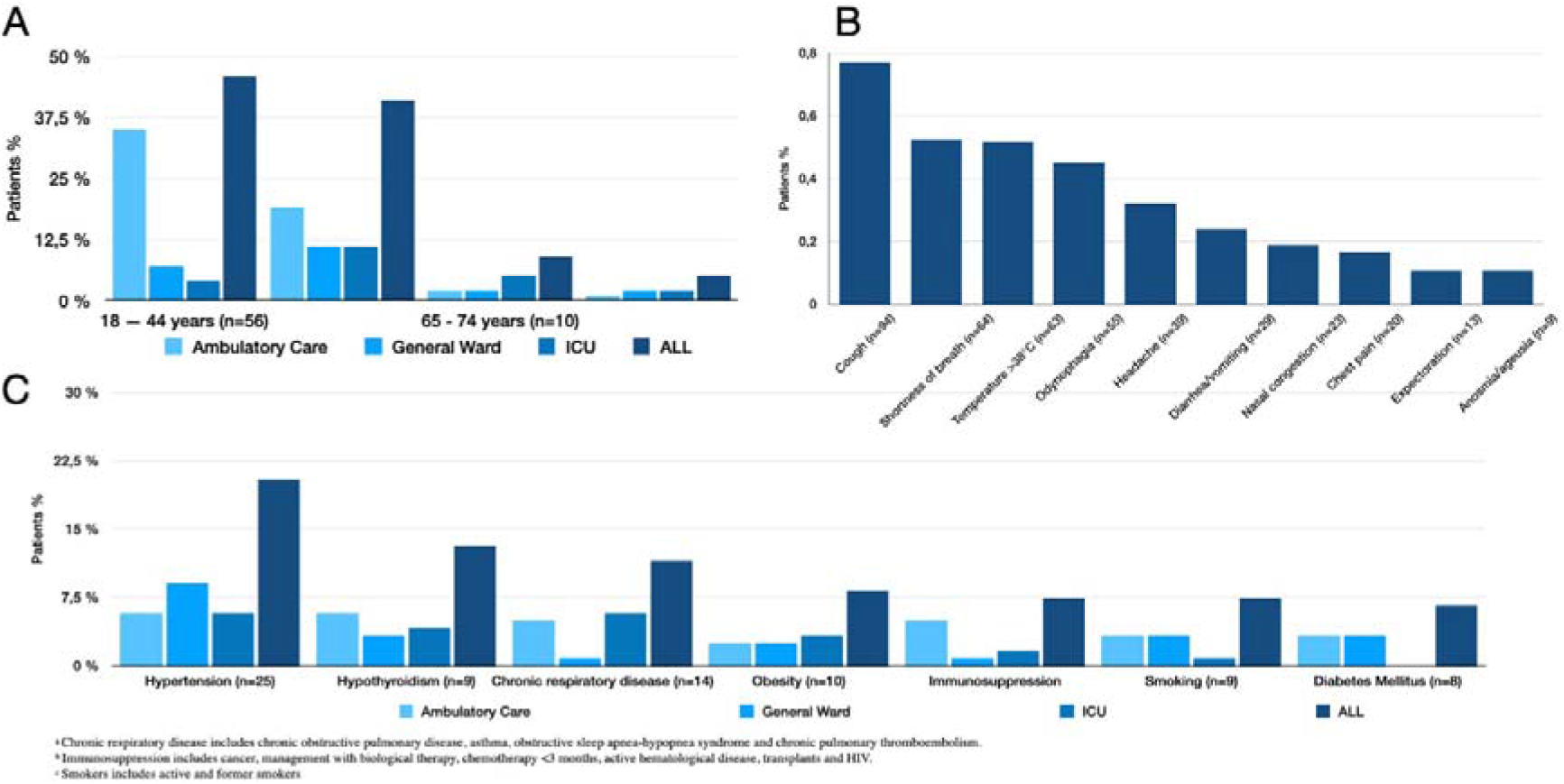
Distribution of age(A), (B)presenting symptoms and (C)comorbidities of adults with SARS-CoV-2, stratified by management setting, in Two Hospitals, Bogota, Colombia

**Table 1.**
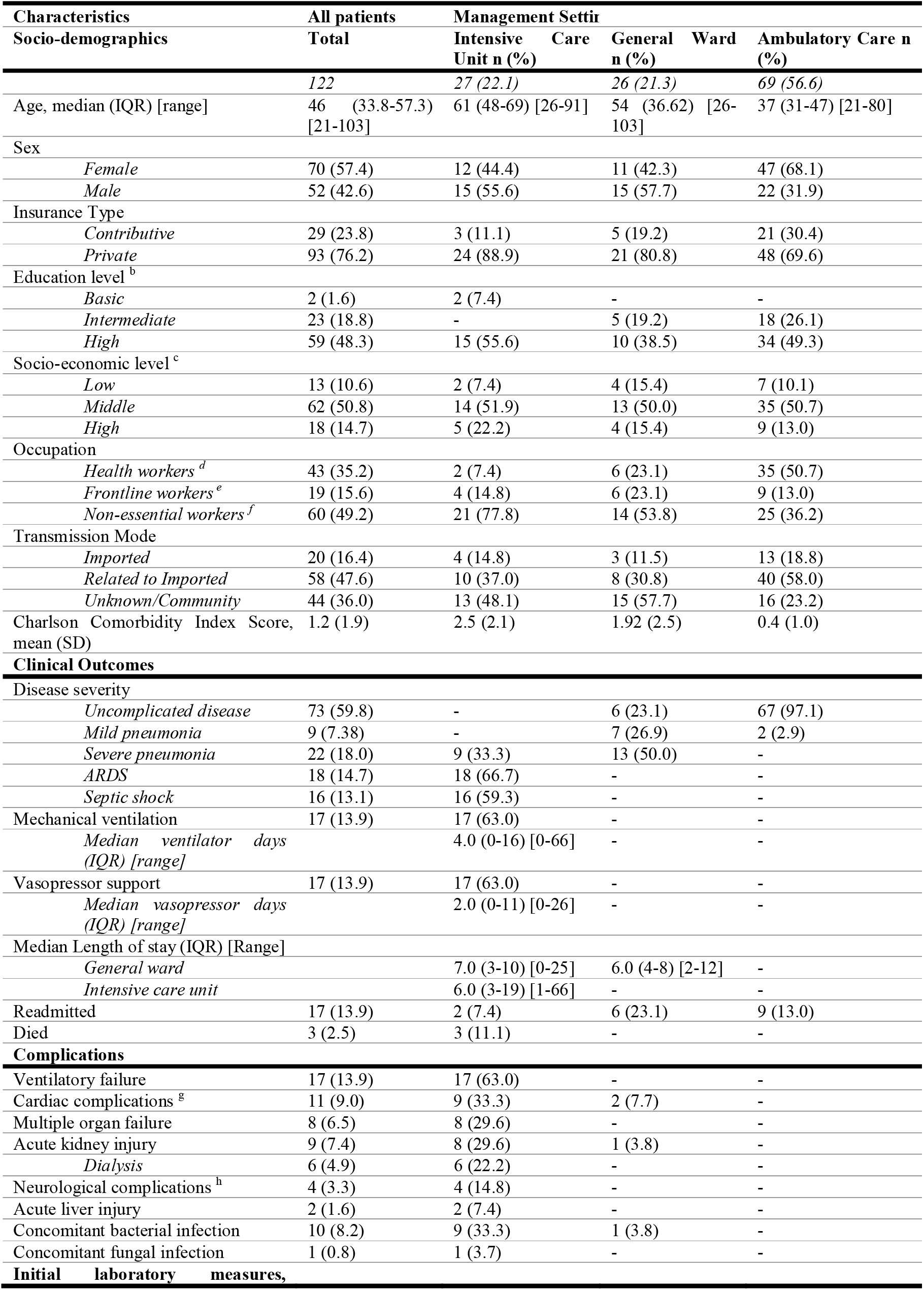
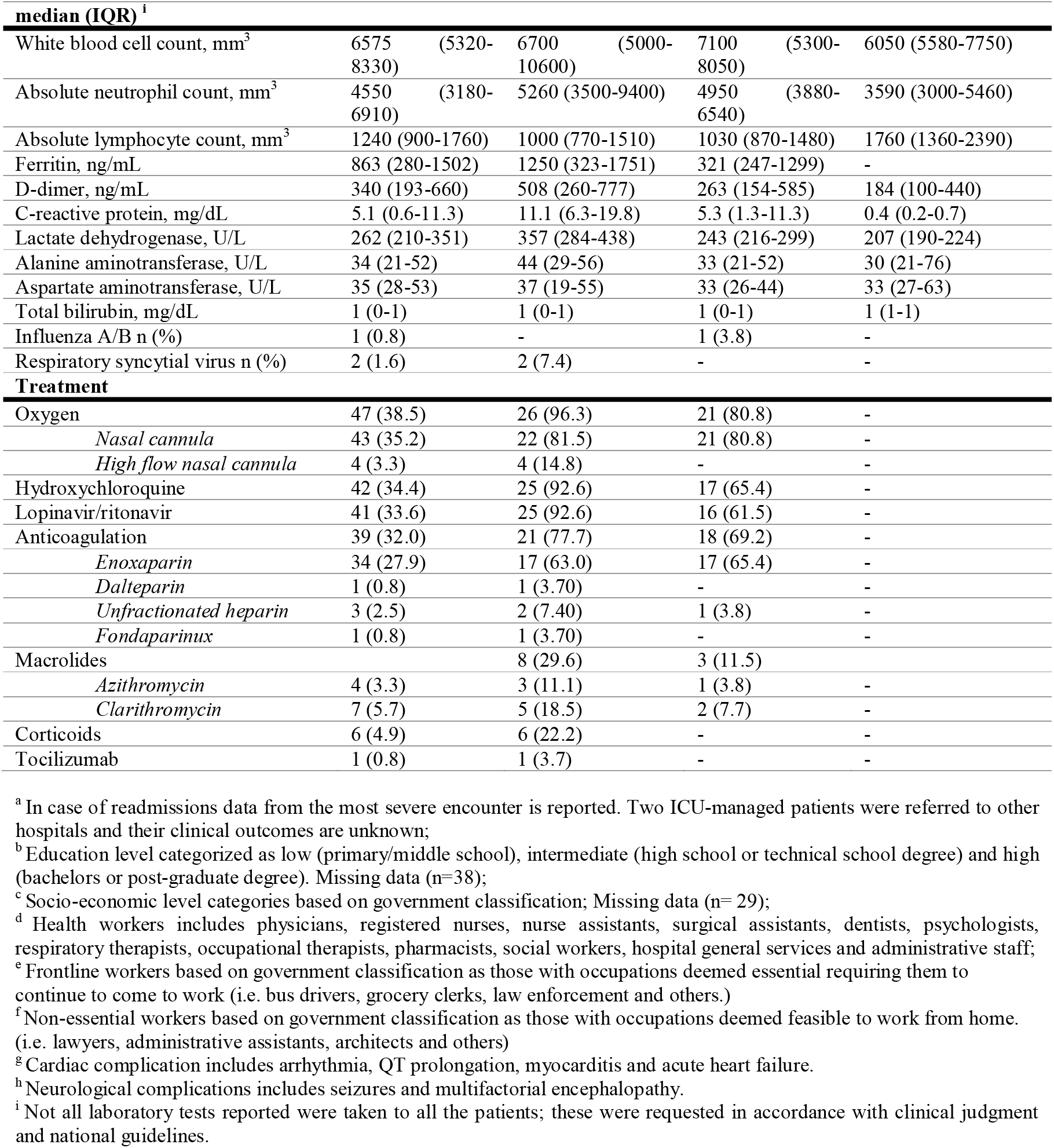
Characteristics of Adults with SARS-CoV-2 Managed in Two Hospitals, Bogota, Colombia^a^

Of the GW patients, 21 (80.8%) required oxygen support by nasal cannula. The primary reason for ICU admission was ventilatory failure with a median length of stay of 6 (3-19) days and 17 (63%) requiring invasive mechanical ventilation.

## DISCUSSION

To our knowledge, this is the first case series of patients with SARS-CoV-2 infection managed in outpatient and inpatient settings in Colombia and LA. With a median age of 46 years, our cohort is younger compared to series from China, Italy and the US.^3,4,5,6^ This may be explained by the population age structure in Bogota and the early adoption of mitigation and suppression strategies, particularly protecting populations 70 and older. The median age and prevalence of comorbidities in our ICU-managed patients were similar to U.S series, with a modest mortality rate (11.1%).

The major limitation of this study is that our series mostly represents the earliest wave of SARS-CoV-2imported infections into the country, primarily among individuals of mid to high SES status. In our cohort, most patients 68 (64%) had a recent history of international travel or were related to imported cases. As the Bogota epidemic achieved sustained community transmission, subsequent infection waves have affected more frontline workers and lower SES status individuals. This is not surprising given the high levels of social inequality in large LA cities,^1^ highlighting the importance of sustained local public health measures to reduce virus transmission and control the overall COVID-19 disease burden in the region.

## Data Availability

I have all the data and its available for further reviews

https://bit.ly/36VE9Sm

## Author Contributions

Drs Moscoso, Sanchez and Rodriguez had full access to all the data in the study and take responsibility for the integrity of the data and the accuracy of the data analysis.

*Concept and design:* All authors.

*Acquisition, analysis, or interpretation of data:* Aya, Moscoso, Sanchez, Lobelo

*Drafting of the manuscript:* Moscoso, Sanchez, Gomez

*Critical revision of the manuscript for important intellectual content:* Rodriguez, Garzon, Lobelo

*Statistical analysis:* Sanchez and Moscoso

*Obtained funding:* N/A

*Administrative, technical, or material support:* Moscoso, Sanchez, Aya, Gomez

*Supervision:* Moscoso, Lobelo.

## Acknowledgements

Special thanks to Dr. Santiago López, Medical Vice President CC & CLC and Dr. Alfonso Correa, Education and Research Subdirector CC & CLC for their support to conduct this study.

